# This time is different: model-based evaluation of the implications of SARS-CoV-2 infection kinetics for disease control

**DOI:** 10.1101/2020.08.19.20177550

**Authors:** Kaitlyn E. Johnson, Madison Stoddard, Ryan P. Nolan, Douglas E. White, Natasha Hochberg, Arijit Chakravarty

## Abstract

As the ongoing COVID-19 pandemic passes from an acute to a chronic situation, countries and territories are grappling with the issue of how to reopen safely. The unique kinetics of infectivity of SARS-CoV-2, with its significant presymptomatic transmission, presents an unprecedented challenge to our intuitions. In this context, a generalizable quantitative understanding of the impact of SARS-CoV-2 infectivity on disease control strategies is vital. We used a previously published time-dependent model of SARS-CoV-2 infectivity (He *et al*., 2020) to parameterize an epidemiological model of transmission, which was then used to explore the effect of various disease control measures. Our analysis suggests that using symptom-based isolation alone as a control strategy is ineffective in limiting the spread of COVID-19, in contrast to its effectiveness in other diseases, such as SARS and influenza. Additionally, timeliness of testing and tracing strategies to reduce time to isolation, along with widespread adoption of measures to limit transmission are critical for any containment strategy. Our findings suggest that for symptom-based isolation and testing strategies to be effective, reduced transmission is required, reinforcing the importance of measures to limit transmission. From a public health strategy perspective, our findings lend support to the idea that symptomatic isolation should not form the primary basis for COVID-19 disease control.

## Introduction

The novel coronavirus SARS-CoV-2, and its associated disease COVID-19, represent a historic threat to human health and well-being. In the absence of effective options for prophylaxis and treatment for this disease, non-pharmacological interventions (NPIs) play a critical role in controlling its spread.

At present, a number of NPIs for COVID-19 have been used in various territories and jurisdictions: symptom-based testing, isolation of symptomatic individuals, measures to limit transmission (such as masks, handwashing and social distancing) and molecular tests for the pathogen. While the first three measures are relatively easy to implement, they have been adopted to varying degrees in different settings, based in part on cultural beliefs (Middelburg and Rosendaal, 2020). On the other hand, molecular tests are less easy to implement at a population level due to their high cost and limited availability and can in many cases take several days to return results. At the time of this writing, for example, the average turnaround time from nasal swab to result for COVID-19 tests in the US was 4.1 days (Baum *et al*., 2020), with 21% of tests returned after five days or more of waiting.

COVID-19 is a pleiotropic disease, with a complex and unpredictable presentation (Huang *et al*., 2020; Wang *et al*., 2020). While there are a number of common symptoms for the disease, including fever, fatigue, dry cough, anosmia and diarrhea, many patients present with diverse symptoms including muscle ache, confusion, headache, sore throat, chest pain, nausea and emesis (García, 2020). In extreme cases, some patients have also presented with blood clots, strokes, and heart attacks (Klok *et al*., 2020). In addition, reports of the symptom profile of COVID-19 have changed over time. Early reports emphasized fever, headache and a cough, while more recent reports emphasized anosmia and diarrhea as major symptoms. As a practical matter, this diversity of symptoms can make it challenging for patients experiencing symptoms of COVID-19 to recognize that they have the disease.

Further complicating disease containment is the biology of SARS-CoV-2 infection. While the initial site of infection for most patients is in the nasopharynx, in more serious cases the infection makes its way to the lungs. Asymptomatic SARS-CoV-2 infections are typically limited to the nasopharynx, with viral loads that are similar to symptomatic infections and decrease more slowly (Lee *et al*., 2020). Presymptomatic transmission is another concern. It is now well established that the peak of infectiousness for COVID-19 occurs about two days before the appearance of symptoms (He *et al*., 2020). Data from contact tracing studies and model-based estimates demonstrate that almost half of all transmission occurs from presymptomatic and asymptomatic patients (Du *et al*., 2020; He *et al*., 2020; Kerr *et al*., 2020; Moghadas *et al*., 2020; Wei *et al*., 2020). Clearly, the stealthy nature of COVID-19 transmission poses a significant challenge for disease-control interventions.

While a number of studies have looked at intervention strategies in specific scenarios (Davis *et al*., 2020; Kerr *et al*., 2020; Tian *et al*., 2020), there is a need for a quantitative investigation that can be applied broadly to questions of disease control strategy. To better understand the impact of SARS-CoV-2 transmission on COVID-19 disease control, we used a time-dependent model of SARS-CoV-2 infectivity. This model was based on infector-infectee pairs that were fitted to a curve to infer the relative infectiousness as a function of time from symptom onset (He *et al*., 2020). We employed the time-variant kinetics of infectivity in a standard susceptible-exposed-infected recovered (SEIR) epidemiological model with two compartments for the infected population (isolated and un-isolated) and examined the impact of various disease control strategies in the light of the kinetics of infectiousness. The time-variant kinetics of infectivity of the 1918 flu and SARS and the resulting impact on disease control are compared to that of SARS-CoV-2 to provide historical context. We hope that these findings shed light on the utility and limitations of various practical real-world strategies for COVID-19 disease control.

## Methods

### SEIR model of SARS-CoV-2 with time-dependent infectiousness

We developed a dynamic transmission model of SARS-CoV-2 infection and assessed the effect of isolating symptomatic individuals on disease transmission under different scenarios. The dynamic, deterministic, compartmental model was based on a SEIR structure, with additional stratification of individuals by whether they were symptomatic (or asymptomatic) and separately whether they were and effectively isolated (Fig. 1A and parameters and references in Table 1). Susceptible individuals (S) can become infected through a dynamic process occurring at a rate proportional to the number of infected individuals and the infectious time course (Fig. 1A). Susceptible individuals (S) become exposed (E) at a rate proportional to the force of infection, λ(t), which is the sum of the number of individuals in each infection stage multiplied by the infectiousness of that stage, given by β(τ) (Fig. 1B). This is the convolution of the number of individuals in each infectious stage and the infectiousness distribution.

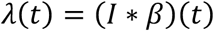

**Figure 1:**
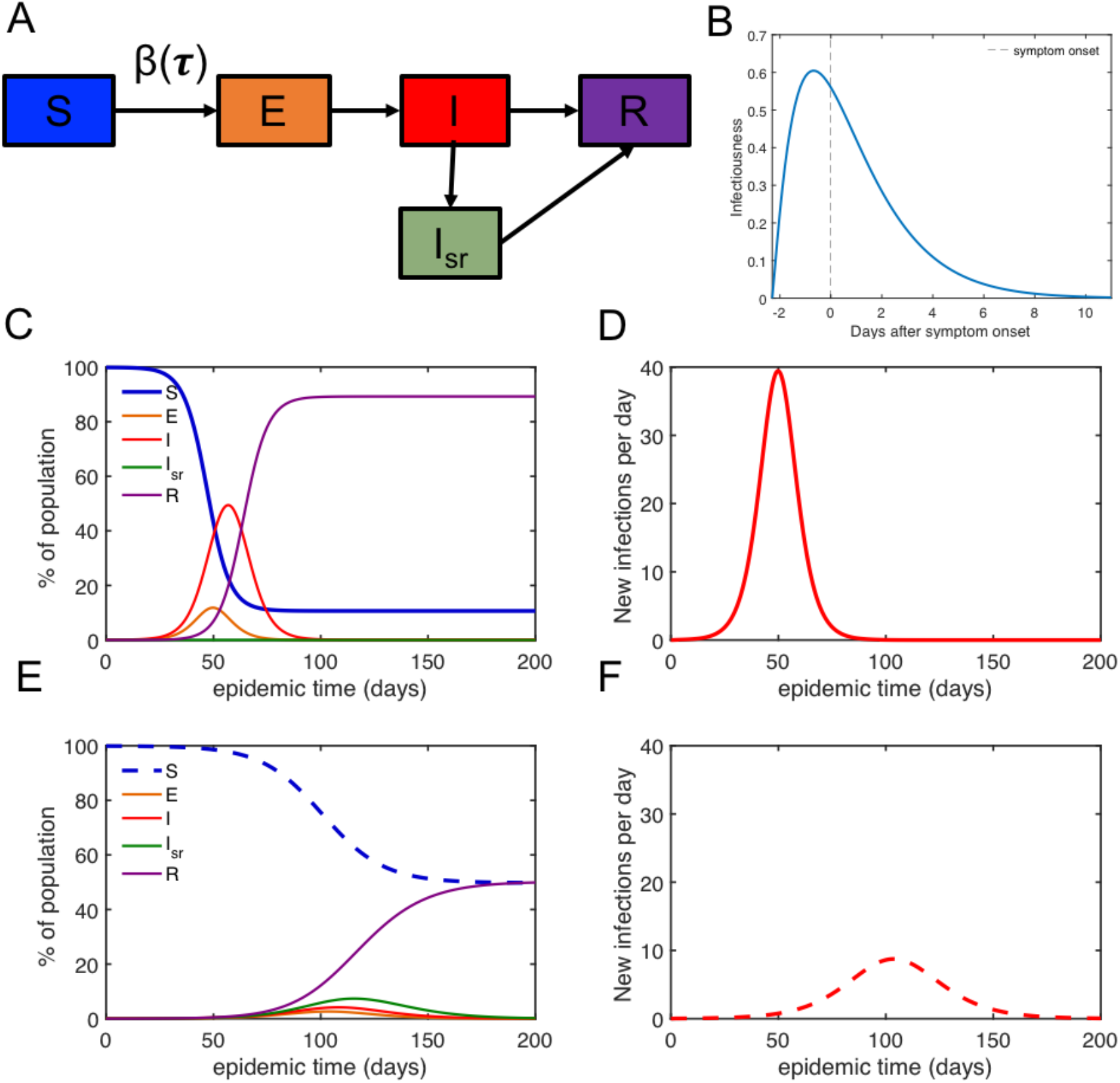
Epidemiological model. A. schematic (SEIR model with isolated and removed compartment) B. Significant pre-symptomatic disease transmission potential for SARS-CoV-2 C. Effect of no isolation strategy on susceptible, exposed, infectious, recovered subpopulations over time, leading to a projected 89% of the population infected. D. Number of new infections per day if no isolation occurs. E. Effect of perfect symptomatic isolation on epidemiological subpopulations over time, leading to a projected 50% of the population becoming infected. F. Number of new infections per day if perfect symptomatic isolation occurs.

The basic reproductive number, R_0_, is the area under the curve of the weighted infectiousness distribution.

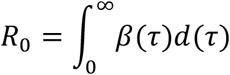

The infectiousness time course is set as a gamma distribution with parameters derived from (He *et al*., 2020), scaled by R_0_. R_0_ represents the average number of infections that an infected individual passes on to others at the initial time point, in the absence of any interventions in a susceptible population. R_0_ was varied throughout this study to simulate changes in the transmission rate of the disease, which depend on NPIs such as hand-washing, mask wearing, and social distancing. The baseline R_0_ for SARS-CoV-2 is conservatively estimated at 2.5 (CDC, 2020a).

Susceptible individuals become exposed (E) but are not infectious for an average of 3 days(Lauer *et al*., 2020). A proportion of the infected individuals (I) develop symptoms after an average of 2.3 days of being infectious (He *et al*., 2020). Upon developing symptoms, individuals seek testing and isolation (I_sr_) at a specific time in their infection stage. We assume that each individual infection is removed at the same time in their infection stage; time to isolation was varied in simulations to evaluate different isolation scenarios. We assume that all infected individuals enter the recovered/removed compartment (R) after 14 days from being infected, where it was assumed that they are immune to reinfection in the time period of interest. The equations for the model are below:

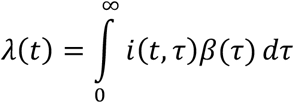

Where i(t,τ) is a matrix containing the number of infectious individuals at each time t in each infection age ***τ***. At each time step dt, the compartments are updated as follows:

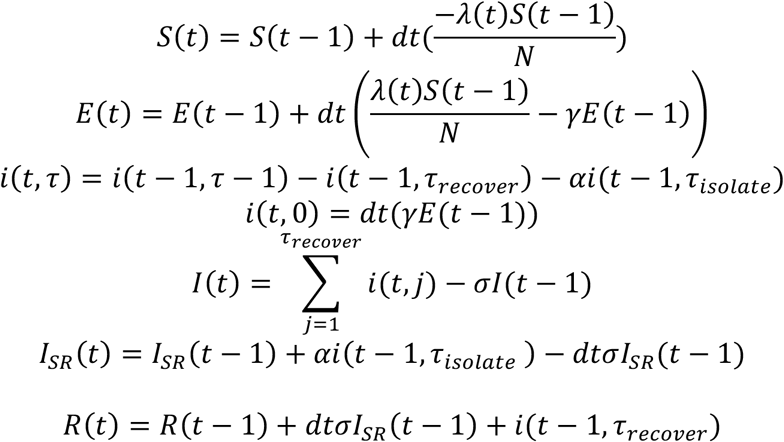

Where N = total population. The initial conditions are I(0) = 1, S(0) =999, R(0)=E(0)=I_sr_(0)=0, for the majority of the simulations. The model parameters and their references are described in Table 1.

**Table 1.** Model Parameters

| Parameter | Symbol | Value | Reference |
| --- | --- | --- | --- |
| Basic reproductive number | $R_0$ | SARS-CoV-2: 2.5 individuals<br>SARS: 3 individuals<br>1918 Flu: 2.5 individuals | (CDC, 2020a)<br>(Pitzer, Leung and Lipsitch, 2007)<br>(Vynnycky, Trindall and Mangtani, 2007;<br>Andreasen, Viboud and Simonsen, 2008) |
| Duration exposed but not detectable or infectious | $1/\gamma$ | SARS-CoV-2: 3 days<br>SARS: 4.5 days<br>1918 Flu: 1.6 days | (Lauer *et al.*, 2020)<br>(Peiris *et al.*, 2003)<br>(Ip *et al.*, 2017) |
| Proportion symptomatic who eventually isolate | $\alpha$ | SARS-CoV-2: varied, max 80%<br>SARS: varied, max 100%<br>Flu: varied max 76% | (CDC, 2020a)<br>(Peiris *et al.*, 2003)<br>(Ip *et al.*, 2017) |
| Duration of illness | $\tau_{\text{recover}}$ | SARS-CoV-2: 14 days<br>SARS: 20 days<br>1918 Flu: | (He *et al.*, 2020)<br>(Peiris *et al.*, 2003)<br>(Ip *et al.*, 2017) |
| Time from initial infection to symptom onset | $\tau_{\text{sym}}$ | SARS-CoV-2: 2.3 days<br>SARS: 0 days<br>1918 Flu: 1.6 days | (He *et al.*, 2020)<br>(Peiris *et al.*, 2003)<br>(Ip *et al.*, 2017) |
| Duration spent infectious before isolating | $\tau_{\text{isolate}}$ | Varies from 1.5 days before symptom onset to 4 days after symptom onset | Varied in simulations |
| Duration spent in isolation before recovering | $1/\sigma = \tau_{\text{recover}} - \tau_{\text{isolate}}$ | Varies, duration of infection (days) – time already spent infectious (days) | Varied in simulations |

### SEIR models of SARS and 1918 influenza with time-dependent infectiousness

We extended the analysis to include analogous SEIR models of both the SARS outbreak in 2003 (Peiris *et al*., 2003) and the 1918 influenza outbreak (Vynnycky, Trindall and Mangtani, 2007; Andreasen, Viboud and Simonsen, 2008). Infectiousness distribution for both SARS (Peiris *et al*., 2003) and flu (Cori *et al*., 2012; Ip *et al*., 2017) were estimated as follows. SARS individuals are not infectious until symptom onset and infectiousness peaks 10 days after symptom onset. We assumed few cases of SARS were subclinical (5% to be conservative). For the 1918 flu, infectiousness begins an average of 1.6 days before symptom onset and peaks 1 day after symptom onset (Ip *et al*., 2017; He *et al*., 2020). Because we are modeling the 1918 flu, we assumed that no individuals were initially immune, just as none are assumed immune for SARS-CoV-2 or SARS, despite some potential role of cross immunity from other flus or coronaviruses. Using estimates from more recent flu seasons that 24% of flu cases are pauci-symptomatic or asymptomatic (Ip *et al*., 2017), we assumed that it was only possible to isolate up to 76% of flu cases using symptom-based isolation alone. The remaining epidemiological parameters and their references are listed in Table 1.

Using the epidemic model, we simulated outbreaks of each disease. We assumed that no infectious individuals enter into the system. The simulations predict the number of individuals in each compartment over time, the new infections per day, the total percent of the population expected to be infected, and the effective reproductive number.

### Estimation of effective reproductive number (R_eff_) for each isolation strategy

The value of R_eff_ was numerically estimated by running a single generation simulation in which 1 infected individual was placed at the first stage of infection in a population of N-1 susceptible individuals. The total number of secondary infections, as the single infection passes through the stages of infection, was recorded and used to directly estimate R_eff_.

### Evaluation of real-world limitations on disease control

To evaluate the effect of *imperfect symptomatic isolation*, we independently varied both the time to isolate after symptom onset and the percent of infections that are symptomatic and eventually isolate. To assess the impact of *delayed isolation*, we increased the time to isolate individuals incrementally, holding the percent symptomatic and eventually removed constant at the “perfect” level (80% for SARS-CoV-2), and recorded the percent not infected, the daily new infections, and the effective reproductive number. To assess the impact of *a lower percent of infections being symptomatic and eventually removed*, we decreased the percent removed incrementally, holding the time to remove individuals constant at the “perfect” level (2.3 days after symptom onset for SARS-CoV-2), and recorded the same outputs as mentioned above. To compare directly the effect of *perfect, realistic, and no isolation strategy*, we simulated a perfect isolation strategy as 80% symptomatic and removed after 2.3 days of infection (immediately after symptom onset), a realistic isolation strategy as 60% symptomatic (CDC, 2020a) and removed after 4.3 days of infection (2 days after symptom onset, (CDC, 2020a)), and no isolation strategy as 0% removed. We recorded the outputs of percent not infected, daily new infections, and the effective reproductive number for each scenario, for SARS-CoV-2, SARS, and 1918 influenza.

### Identify regions of parameter space that enable outbreak control

To identify regions of parameter space that enable outbreak control, we varied simultaneously the percent of the population eventually removed and the time to remove infectious individuals, and we recorded the total percent infected and the effective reproductive number. For this analysis, the time to remove infectious individuals was anytime during the infection, which for SARS-CoV-2 and flu, this could be before symptom onset. This was meant to simulate the theoretical possibility that individuals could be isolated based on molecular testing or contact-tracing prior to symptom onset. This set of simulations was performed over a range of transmission rates corresponding to 50%, 75%, 100%, and 150% of the R_0_ used in the base simulations, in order to simulate the effect of transmission reduction measures such as mask wearing and social distancing, and as well as to simulate high contact scenarios such as a university. This was performed for SARS-CoV-2, SARS, and 1918 influenza. The analysis of strategy space as a function of transmission reduction and time to isolation uses a realistic assumption of the percent of cases that were symptomatic (60% (Kerr *et al*., 2020)).

### Critical basic reproductive number to control outbreak

To determine the baseline transmission rate needed to keep the outbreak under a critical threshold using symptomatic isolation alone as a mitigation strategy, we varied the basic reproductive number, R_0_, and recorded the expected total percent infected and effective reproductive number for no isolation (α = 0), realistic symptomatic isolation (***τ***_isolate_ = 4.3 days, 2 days after symptom onset, α = 60%, and perfect symptomatic isolation(***τ***_isolate_ = 2.3 days, immediately upon symptom onset, α = 80%). We repeated this analysis assuming that testing strategies enable isolation of infectious individuals at 1.5, 1, and 0 days before symptom onset. The threshold for total percent infected was defined somewhat arbitrarily at twenty percent, based on the estimated proportion of New York City that was infected in March-April 2020 (Stadlbauer *et al*., 2020). The critical threshold for the effective reproductive number was chosen as one, as this would mean that effectively the outbreak would be contained, mitigating significant future spread.

## Results

### Evaluating the effectiveness of symptomatic isolation as a containment strategy

Our analysis reveals that perfect symptomatic isolation compared to no isolation measures reduces the expected total percent infected from 89% to 50% (Fig 1 C-F), indicating that perfect symptomatic isolation can have a significant impact on control of SARS-CoV-2. Because symptom onset can be slow and variable (Huang *et al*., 2020; Wang *et al*., 2020), the effect of a delay in symptomatic isolation was evaluated, revealing that even a one-day delay in isolation leads to a significant increase in the total percent of the population infected (Fig. 2 A-C). Additionally, the estimated percent of asymptomatic individuals varies, from 20-40% for the general population and even higher for younger populations. When the percent of the infections removed is lowered, the outbreak worsens (Fig. 2 D-F). We compared the “realistic” symptomatic isolation strategy to the “perfect” and “no isolation” strategy and we see that the realistic strategy is much less effective at reducing the total of the population infected, the peak daily rate of new infections, and the effective reproductive number (Fig. 2 G-I). The realistic strategy appears to follow a similar trajectory of transmission to no isolation at all, indicating that slight relaxations from perfect symptomatic isolation render the strategy much less effective at mitigating spread of disease.

**Figure 2:**
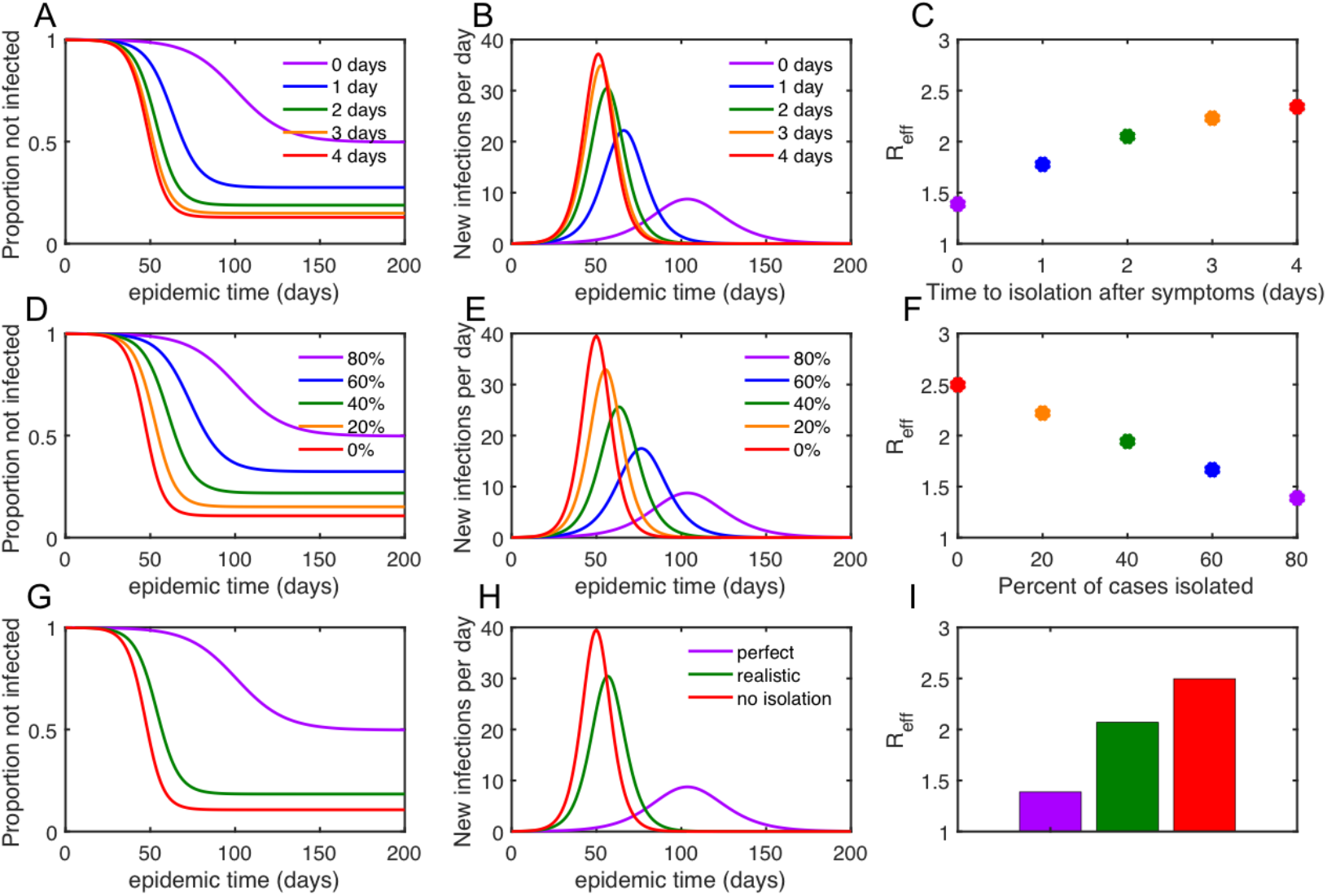
Impact of real-world limitations on the effectiveness of disease control for SARS-CoV-2. A-C. Effect of the number of days post-symptom onset that infectious individuals are isolated. D-F. Effect of the percent of the cases able to be isolated. G-H. Comparison of a perfect symptomatic isolation strategy, a realistic symptomatic isolation strategy, and no isolation strategy at all, which shows that for realistic limitations on disease control, the effectiveness of symptomatic isolation is close to doing nothing at all.

### Exploring the effectiveness of timeliness of isolation, degree of adoption, and rate of disease transmission on outbreak control

Varying the time to isolation and the degree of adoption of isolation, the regions of parameter space that enable a reasonable control of an outbreak (R_eff_<1, white line) do not fall within the region of symptomatic isolation alone strategy (green box, Fig. 3C&G) for the R_0_ of SARS-CoV-2 (conservatively estimated as 2.5). As the transmission rate (R_0_) increases, this becomes even more improbable, with a high percent of the infections needing to be isolated more than a full day before symptom onset for outbreak control (Fig. 3 D&H). However, as the R_0_ decreases, symptomatic isolation alone as a strategy becomes more feasible, with a 50% reduction in transmission resulting in a significant portion of reasonable strategy space enabling an R_eff_ less than 1 (Fig. 3A&E). The initial number of infections does not affect the expected total percent infected as a function of parameter space (Fig. S1), with a lower percent initially infected corresponding to a longer time-to-peak new infections (Fig. S2). Without lowering the transmission rate, SARS-CoV-2 is impossible to contain via symptomatic isolation. However, depending on the efficacy of the isolation strategy, small changes in transmission can have a significant effect on the total percent of the population infected.

**Figure 3:**
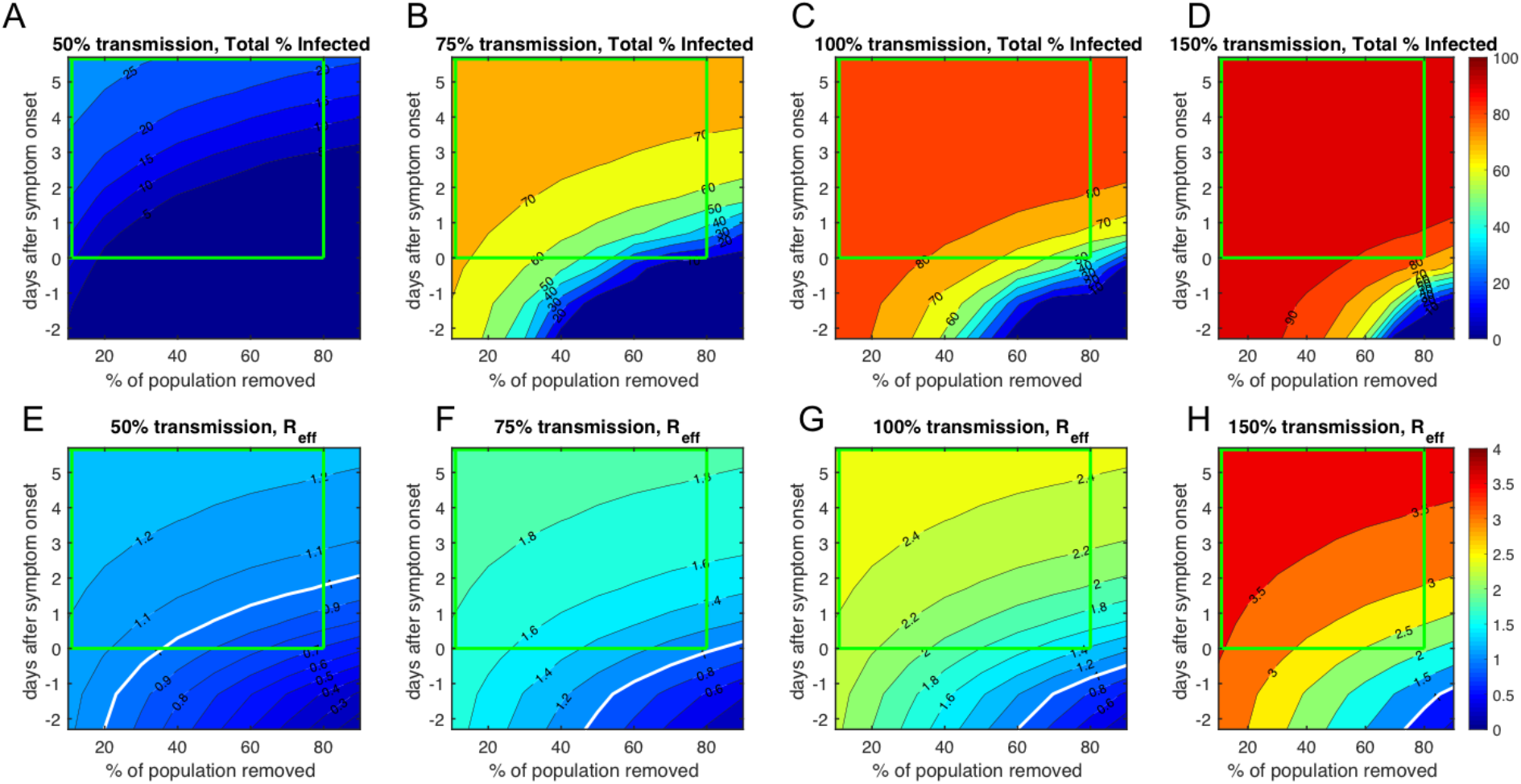
Regions of parameter space that enable outbreak control of SARS-CoV-2. A-D: Projected total percent infected under transmission rates of 50%, 75%, 100%, and 150% relative to baseline, representing R_0_s of 1.25, 1.875, 2.5, and 3.75. The percent of infectious individuals able to be isolated and the timeliness of isolation are varied. Green box outlines the feasible region of parameter space under symptomatic isolation. E-H. Effective reproductive number (R_eff_) under the same parameter sweeps as in A-D, with green box representing feasible region of symptomatic isolation strategy. White lines represent the boundary of R_eff_=1, where below this boundary the outbreak can be controlled. The analysis reveals that for a COVID R_0_ of 2.5 (100% transmission), symptomatic isolation is not effective at bringing the R_eff_ below 1, but at reduced transmission levels the strategy can be effective.

For a given isolation strategy (perfect, realistic, and no isolation), our analysis reveals the effect of reducing transmission (R_0_) on the total percent infected (Fig. 4). We define a threshold of 20% total infected to show an example of the critical R_0_ needed for each strategy. Our analysis reveals that if no isolation is expected to occur, the transmission needs to be reduced by greater than 50% (R_0_ = 1.2), if a realistic symptomatic isolation strategy is in place, transmission needs to be reduced by 40% transmission (R_0_ = 1.5), and if perfect symptomatic isolation is in place, transmission needs to be reduced by 12% (R_0_ = 2.2). The non-linearity of the relationship between total infections and transmission indicates that, for a specific strategy, small reductions in transmission can have a significant effect on reducing the number of infections. None of the symptomatic isolation strategies are effective at reducing total infections to below 20% of the population with the baseline transmission of SARS-CoV-2.

**Figure 4:**
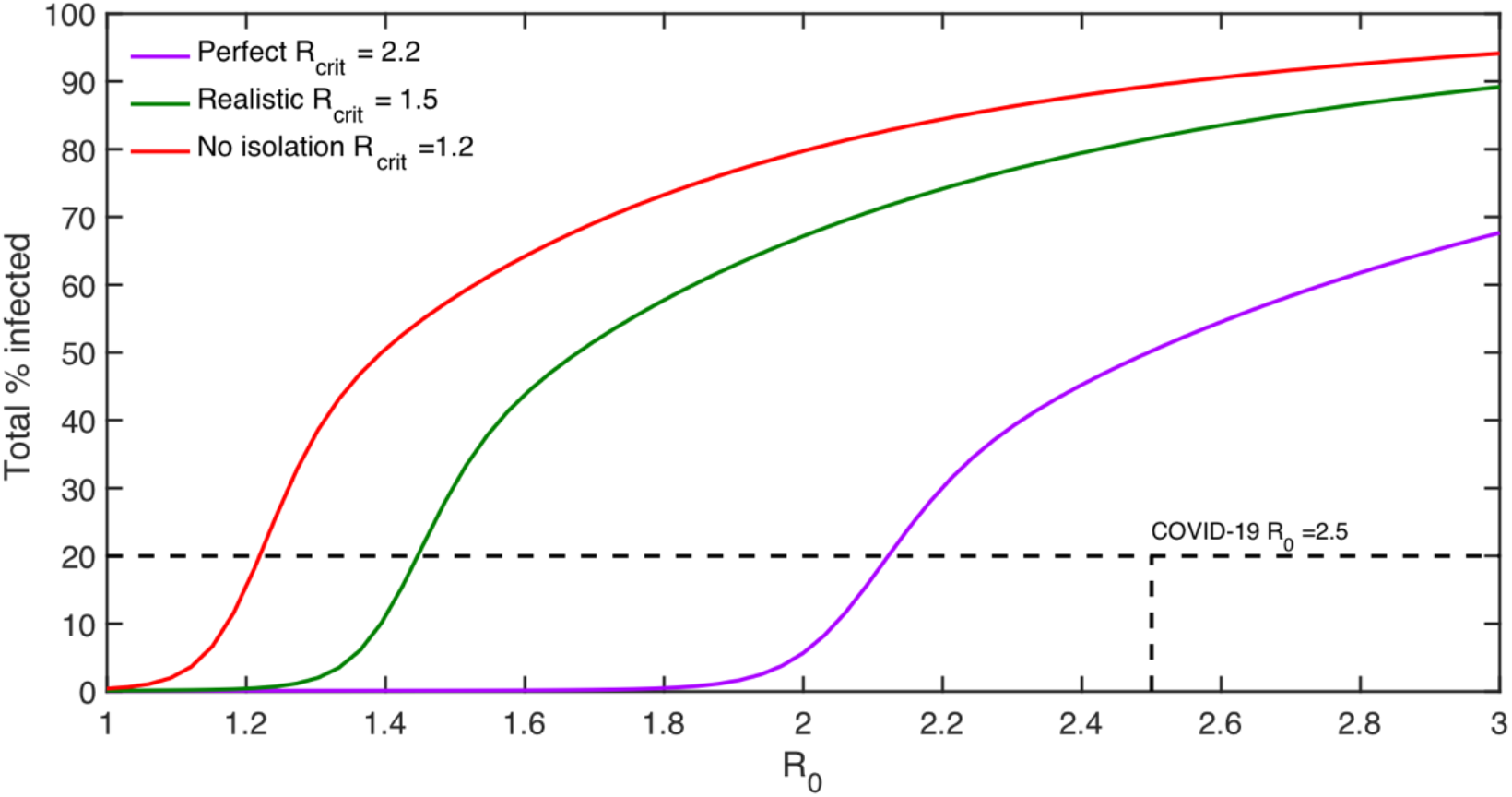
Critical R_0_ for symptomatic isolation of SARS-CoV-2. Total percent infected during the epidemic as a function of the baseline transmission (R_0_) under perfect, realistic, and no symptomatic isolation strategies. For all symptomatic isolation strategies, the R_0_ must be reduced from its estimated baseline of 2.5 in order to mitigate the total percent infected from crossing above 20%.

### Comparing SARS-CoV-2 to SARS and the 1918 influenza, both were more easily contained via symptomatic isolation

SARS symptoms develop before infectiousness begins, and peak 10 days after symptom onset (Fig S3A), and few asymptomatic cases were reported (Ip *et al*., 2017). The regions of parameter space for effective symptomatic isolation for SARS are therefore much larger, and infections can be easily contained even with significant relaxations from perfect symptomatic isolation and increased transmission (Fig. 5, Fig. S3B-E, Fig. S4).

**Figure 5:**
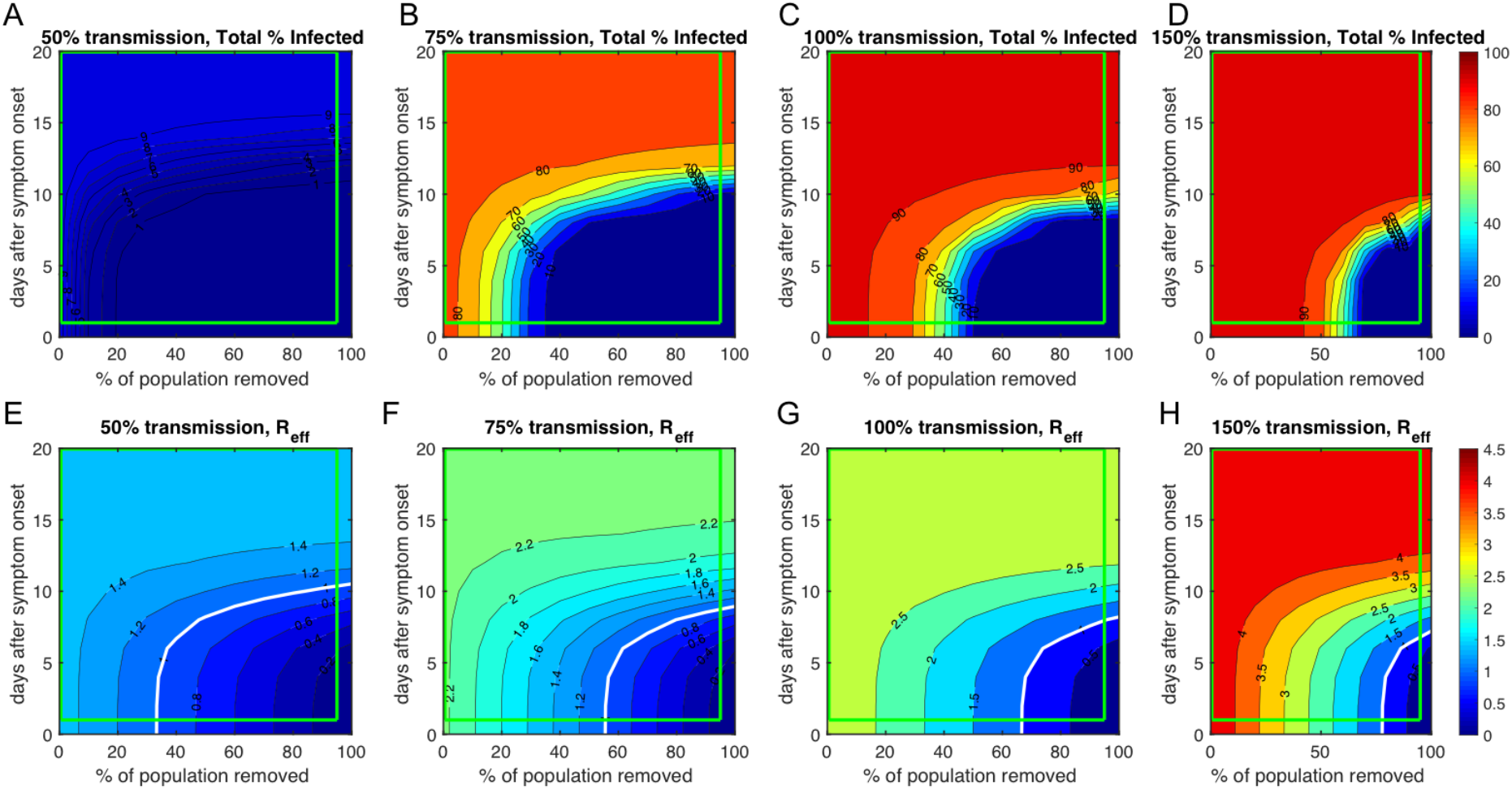
Regions of parameter space that enable outbreak control of SARS. A-D: Projected total percent infected under transmission mitigation of 50%, 75%, 100%, and 150%, representing R_0_s of 1.5, 2.25, 3, and 4.5 for combinations of the percent of infectious individuals able to be isolated and the timeliness of isolation. Green box outlines the feasible region of parameter space under symptomatic isolation. E-H. Effective reproductive number (R_eff_) under the same parameter sweeps as in A-D, with green box representing feasible region of symptomatic isolation strategy. White lines represent the boundary of R_eff_=1, where below this boundary the outbreak can be controlled. The analysis reveals that for SARS, symptomatic isolation was effective at bringing the R_eff_ below 1, even for higher than normal transmission levels.

The 1918 influenza, in contrast, is similar to SARS-CoV-2. Based on data collected from recent flu strains, infectiousness begins 1.6 days before symptom onset, but peaks 1 day after symptom onset (Peiris *et al*., 2003) (Fig. S5A) and up to 24% of cases may be asymptomatic or paucisymptomatic (Ip *et al*., 2017). While these numbers come from recent flu strains, we expect them to be relatively consistent across flu strains. Our analysis reveals that at the baseline R_0_ of the 1918 flu, symptomatic isolation as a control strategy is possible, but only if it is perfect (Fig. 5G, Fig. S5, Fig S6), as is shown in Figure 5G where the green line represents symptomatic isolation strategy and the white line defines part of the boundary between R_eff_ <1 and R_eff_>1. Containment via symptomatic isolation becomes easier if the transmission is reduced by 75% (Fig. 6A, B, E, F), and impossible if the transmission is increased (Fig. 6 D, H).

**Figure 6:**
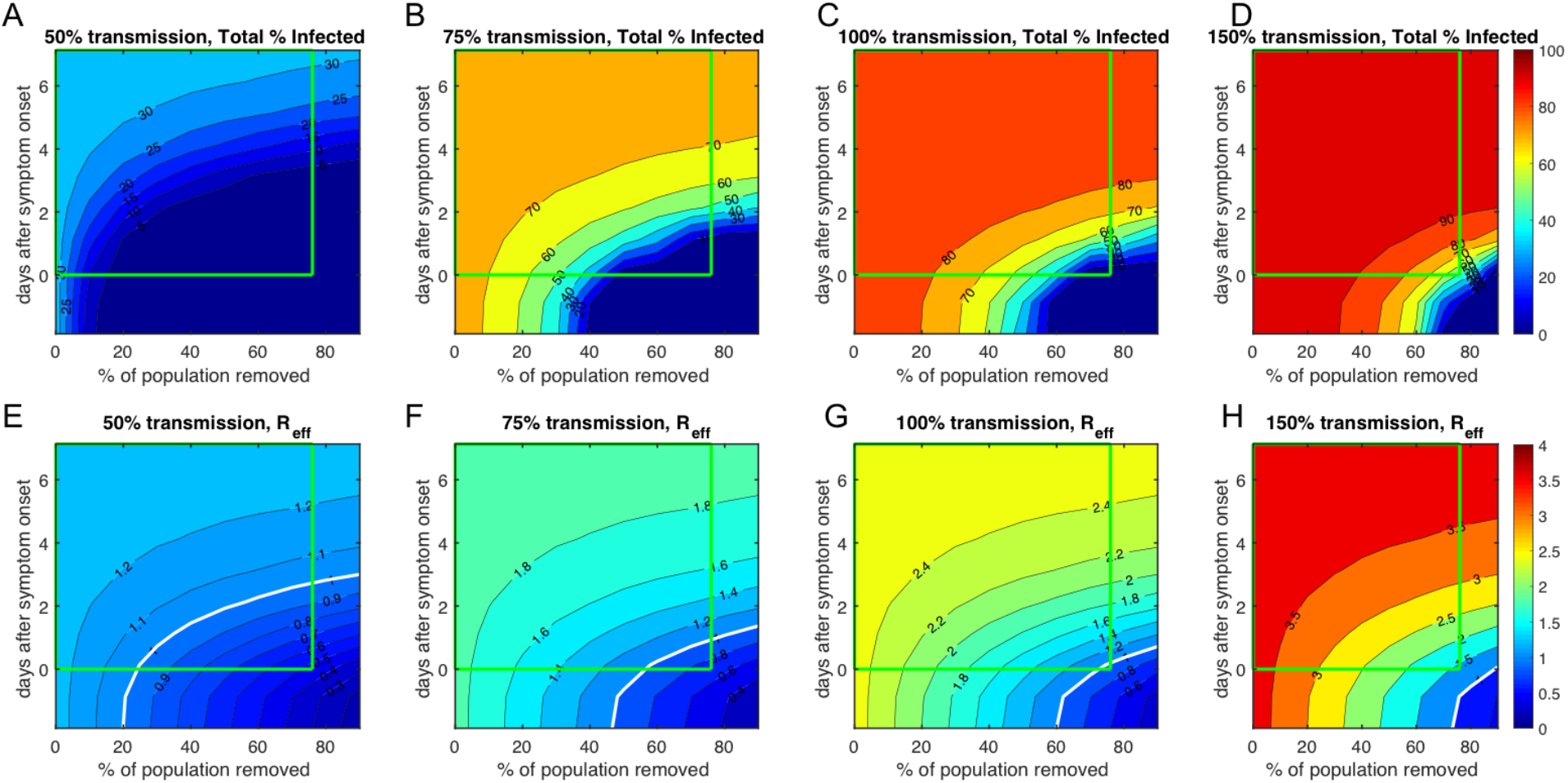
Regions of parameter space that enable outbreak control of 1918 influenza. A-D: Projected total percent infected under transmission mitigation of 50%, 75%, 100%, and 150%, representing R_0_s of 1.25, 1.875, 2.5, and 3.75 for combinations of the percent of infectious individuals able to be isolated and the timeliness of isolation. Green box outlines the feasible region of parameter space under symptomatic isolation. E-H. Effective reproductive number (R_eff_) under the same parameter sweeps as in A-D, with green box representing feasible region of symptomatic isolation strategy. White lines represent the boundary of R_eff_=1, where below this boundary the outbreak can be controlled. The analysis reveals that for an influenza R_0_ of 2.5 (100% transmission), symptomatic isolation is just barely able to bring the R_eff_ below 1.

### Rapid testing and transmission reduction measures are needed to bring a SARS-CoV-2 outbreak under control

We extended our analysis to allow for isolation prior to symptom onset, theoretically achieved through rapid and widely available testing. Here we assumed a constant 60% of cases were isolated through any means. If testing strategies can reduce the average time to isolation, then smaller reductions in transmission are required for outbreak containment below the 20% threshold. Transmission must be reduced by 28% (R_0_ = 1.8) for isolation immediately at symptom onset and 8% (R_0_ = 2.3) for isolation 1 day before symptom onset (Fig. 7) to bring the outbreak below a 20% total infection rate. Isolation 1.5 days prior to symptom onset represents the only scenario where the outbreak can be controlled without reduction in transmission (assuming a COVID R_0_ = 2.5). None of the symptomatic isolation strategies are effective at reducing total infections to below 20% of the population with the baseline transmission of SARS-CoV-2(Fig. 3), but increasing the speed at which infected individuals are isolated can lead to containment under certain transmission reduction scenarios (Fig. 7)

**Figure 7:**
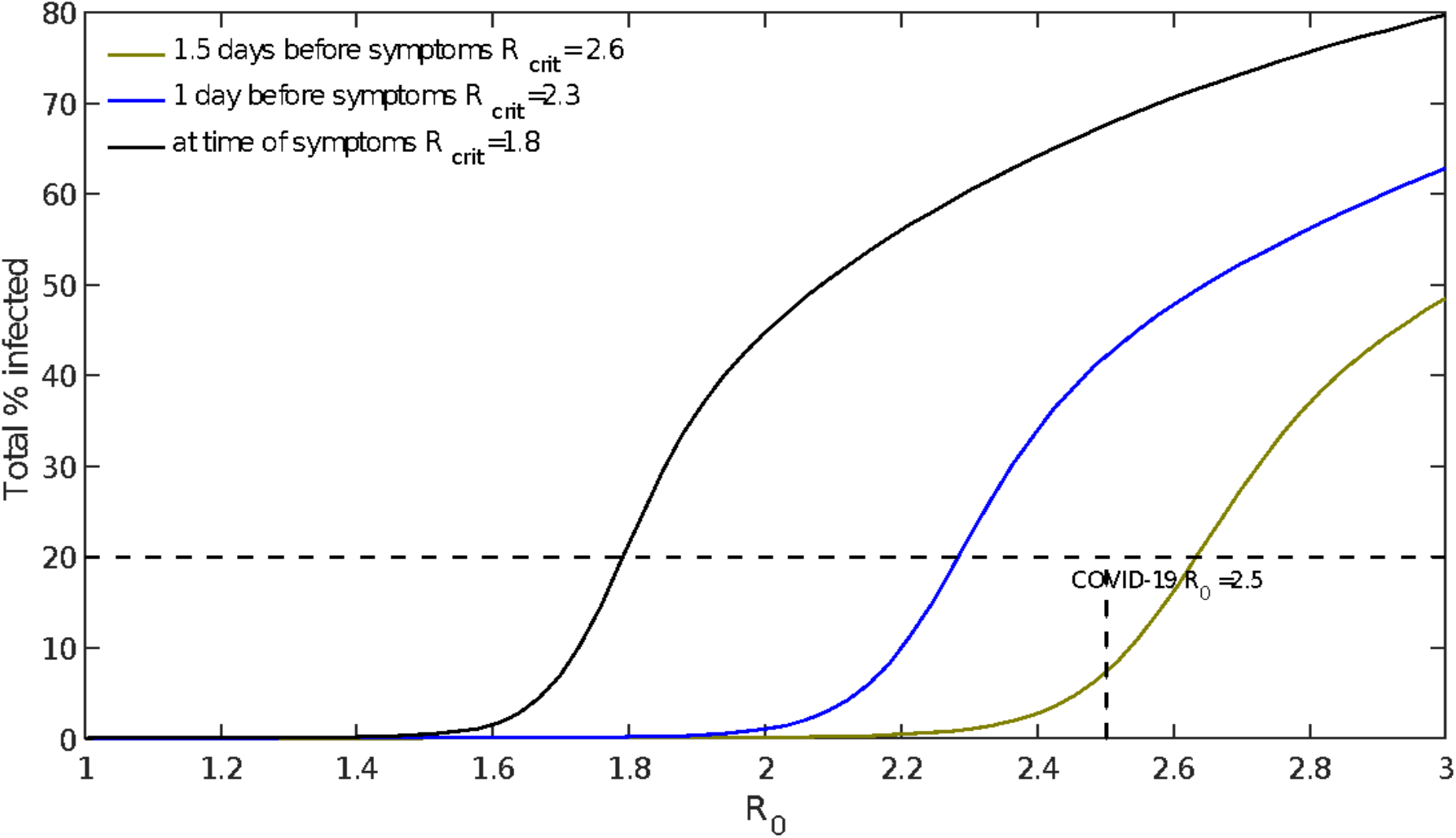
Critical R_0_ for pre-symptomatic isolation of SARS-CoV-2. Total percent infected during the epidemic as a function of the baseline transmission (R_0_) under testing strategies that enable isolation on average 1.5 days before symptom onset, 1 day before symptom onset, and immediately upon symptom onset. For 1 day before symptom onset and immediately upon symptom onset, the R_0_ must be reduced by other means. If the average time of isolation is 1.5 days or more before the onset of symptoms, the R_0_ can remain at or above the COVID R_0_ of 2.5 to prevent more than 20 % of the population from becoming infected.

Simulating increased testing capacity/contact tracing to reduce time to isolation and increased mask use/social distancing, we find that both measures contribute to reducing R_eff_ below 1 (Fig. 8a). We assume that increased capacity, efficiency of testing, and contact tracing would allow access to the region below 0 on the y-axis (Fig. 8a test tube graphic), corresponding to isolation of individuals prior to symptom onset, when they otherwise wouldn’t know they had been infected. If transmission is not reduced (i.e.% transmission = 100), then R_eff_ can only be brought below 1 if isolation occurs prior to symptom onset (Fig. 8b, Fig 8a). The rate of increase in R_eff_ is very steep for the region between 2 days before and 2 days after symptom onset, indicating the potential gain of rapid testing and isolation. Increased mask use and social distancing measures such as avoiding crowded indoor spaces to reduce transmission (Fig. 8 mask graphic) also have a significant effect on bringing the R_eff_ under control. If isolation occurs immediately upon symptom onset, R_eff_ can be reduced below 1 if transmission is reduced by 60% (Fig. 8c, Fig. 8a red dot). This analysis reveals that measures to reduce transmission via social distancing, shutdowns, and mask-wearing can reduce R_eff_ below 1, but to resume normal activities in the absence of significant population immunity, mass rapid testing to shift time to isolation on average to be below time of symptom onset will be needed to enable epidemic control of SARS-CoV-2. Symptom-based testing and isolation alone will inevitably lead to epidemic spread in the absence of significant measures of transmission reduction.

**Figure 8:**
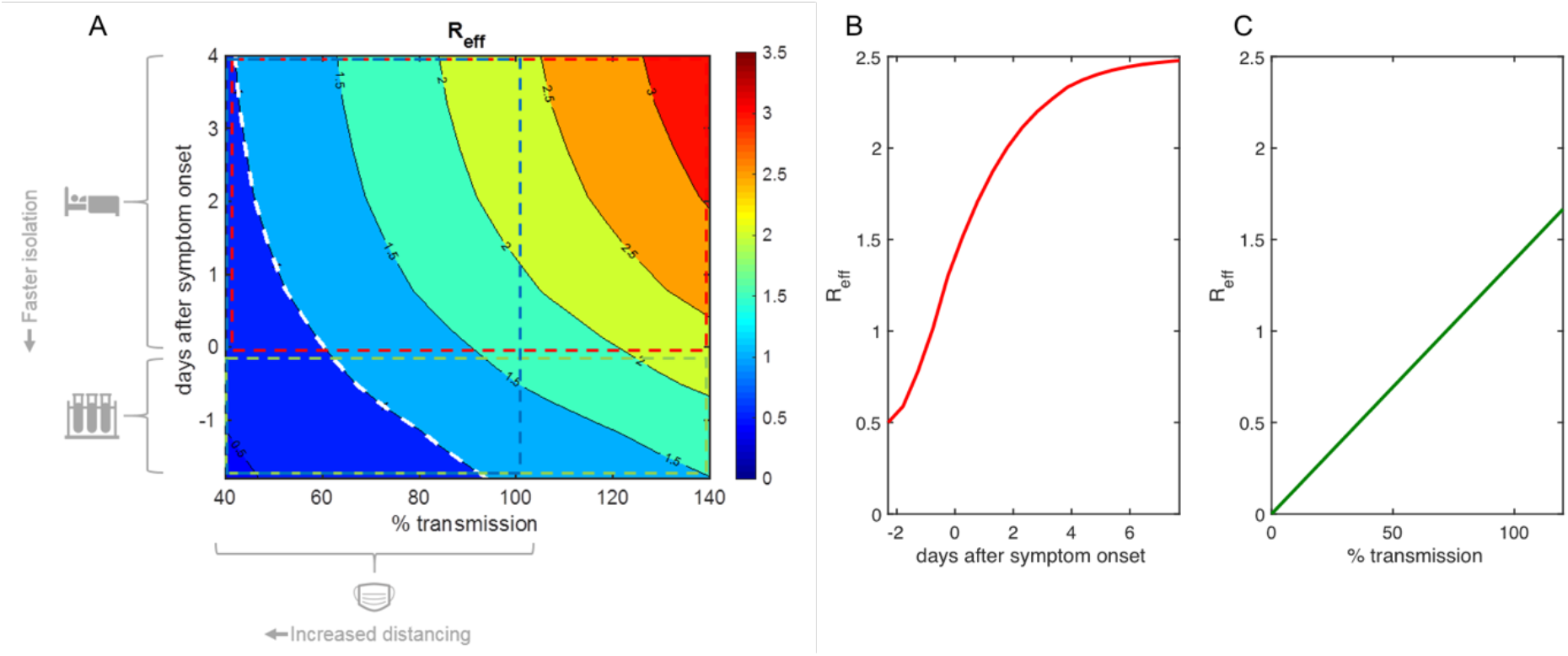
Rapid testing and transmission reduction are both needed to bring outbreaks under control. A. Effective reproductive number (R_eff_) as a function of timeliness of isolation and percent transmission. B. Effective reproductive number (R_eff_) versus timeliness of isolation for 100% of baseline transmission indicates nonlinear effect on increased (R_eff_). C. Effective reproductive number (R_eff_) versus percent transmission indicates that R_eff_ scales linearly with the amount of transmission reduction.

## Discussion

In the early stages of a pandemic, effective control relies on the simultaneous application of multiple NPIs. Mathematical modeling of the 1918 influenza outbreak has shown that early application of multiple simultaneous measures, primarily measures to limit transmission, was associated with a substantial reduction of peak death rates (Hatchett, Mecher and Lipsitch, 2007).

For COVID-19, NPIs can be divided into three kinds of measures: testing, symptomatic isolation and approaches to limit transmission. While it is unlikely that any one measure will be implemented in isolation, resource limitations can sometimes lead organizations and territories to rely more heavily on one measure than others. Therefore, it is important to understand the impact of individual NPI measures separately, in order to tease out the contribution that each measure makes to COVID-19 disease control. In this work, we have used a model-based approach to identify generalizable principles for COVID-19 public health strategy.

Testing for symptoms and isolating individuals is an intuitive and easily implemented strategy for public health authorities during an outbreak or epidemic. The concept of quarantine is an ancient one, with an etymology rooted in the Black Death and references dating back to the Bible (Leviticus 13:4). In recent times, SARS in particular was very effectively controlled through public health interventions. Our work suggests that this may have happened in some measure through symptomatic isolation, although studies have raised questions about the cost-benefit of temperature screening in that context (Chng *et al*., 2004; St John *et al*., 2005; Mouchtouri *et al*., 2019).

For the current pandemic, many territories and organizations have made symptomatic testing and isolation an integral part of their strategy for disease control. For example, Walt Disney World in Florida uses temperature checks on all guests and employees and uses a cutoff of two consecutive temperature readings of 100.4 F to send employees home (Alicia, 2020). Similar strategies are being adopted in other resource-constrained settings, such as public schools, where temperature checks are being adopted as the primary means of controlling the spread of COVID-19. Symptomatic isolation is cheap to implement and has the appeal of transferring the burden of responsibility to individuals, which is particularly important in low-resource situations.

Despite the historic appeal and the ease of implementation, our work shows that symptomatic isolation measures cannot be used as the sole basis for COVID-19 disease control. In fact, symptomatic isolation is only useful to the extent that it is followed up with contact tracing, or other measures that can reduce the time to isolation to at or before the emergence of symptoms. These findings are consistent with other studies on this subject. Model-based analyses show that symptomatic screening for COVID-19 at airports would be expected to miss around 46% of infectious travelers (Quilty *et al*., 2020). News reports from the US from late February indicated that no cases of SARS-CoV-2 had been detected by temperature screening at airports (Cohen and Bonifield, 2020), despite airport screening having been in place (CDC, 2020b) for over a month at that time. During the same timeframe, retrospective reports have demonstrated that community spread was already under way within the country (Jorden *et al*., 2020).

While others have commented on the ineffectiveness of symptomatic testing as “public health theater” (Stanley, 2020), our work suggests that symptomatic isolation is nonetheless valuable as part of a larger strategy, as it will have the effect of limiting spread. In this context, it is important for authorities to err on the side of caution when it comes to interpreting COVID-19 symptoms, and to have generous sick leave policies that enable isolation for those who are infected.

In more general terms, the effectiveness of any testing-and-isolation strategy (whether based on symptoms or molecular markers) is strongly dependent on the speed of implementation of isolation measures (Fig. 8b). Our work suggests that testing-based strategies are particularly useful in the first few days after infection. However, delays of more than a few days degrade the value of test results in a nonlinear fashion, such that at day 4 after symptom emergence there is little to no benefit of test results for the infected patient. In countries where testing is being limited to symptomatic patients, and where there are delays in testing (such as the United States), this limits the benefit of the testing for isolation and contact-tracing. In some jurisdictions, the responsibility of isolation and contact tracing has been shifted on to individuals, making timeliness and compliance an open question (Minnesota Department of Health, 2020).

Our work underscores the importance of measures to limit transmission. These are particularly relevant in the context of reopening strategies-reopening in the presence of high levels of community transmission places a very high burden on testing-and-isolation strategies. On the other hand, reductions in transmission substantially reduce the burden on testing-and-isolation, making it easier to obtain benefit from this measure. Because measures to reduce transmission (such as mask usage) can also be quite cheap and easily implemented, it is imperative to emphasize their importance in public health messaging around COVID-19 disease control. Interestingly, our work shows that the benefit of measures to limit transmission scale linearly with their effectiveness, suggesting that any effort that public health authorities put in to limiting transmission is likely to be rewarded.

We note that there is a point in this figurative “parameter space” that is actually quite practically achievable for public health authorities. Using a test-and-isolate strategy, if the average time of tests returned is around day 0 of symptom onset, and the baseline R_0_ has been reduced to about 60%, the effective rate of spread (R_eff_) is then at or below 1. So, in practical terms, if public health authorities for a given region can bring the rate of community spread down somewhat (corresponding to an R_0_ of about 1.5 for realistic symptomatic isolation, Fig. 4, Fig. 8), surveillance testing strategies with reasonable turnaround times (1-2 days) can be expected to be sufficient to control transmission. This is a feasible strategy for high-resource settings, and it is worth noting that a number of organizations appear to be trying to employ this strategy, as of this writing (Harvard University, 2020; Ward-Henninger and Maloney, 2020).

Viewed in the context of the evolutionary arms race between host and pathogen, pandemics are in some sense a measure of our society’s technological evolution. The bubonic plague was hard or impossible to control before the germ theory of disease, cholera was difficult to control before the development of epidemiology as a science, and influenza was difficult to control before refinements in NPIs and the development of vaccines. In our recent history, we have had examples of outbreaks such as SARS that were adequately controlled by measures and technologies readily available to us.

COVID-19 represents a challenge of a different scale. Viewed in this context, it is easy to understand why this disease in particular has had the ability to grow to a pandemic level. It challenges our intuitions (around the effectiveness of symptomatic isolation, for example), it strains our operational capabilities (in particular with the turnaround time for testing), and it tests our resolve (around the commitment to measures to limit transmission).

Our work shows the importance of a multipronged approach in bringing to heel this particular disease with its unique transmission kinetics, and points to the importance of rapid testing/tracing-based isolation and broader reductions in community transmission.

## Data Availability

No data was used in this work.

## Notes

### Competing Interest Statement

The authors have declared no competing interest.

### Funding Statement

The authors have no funding sources to declare.

